## Supplementary Materials for "Genetic surveillance of *Plasmodium falciparum* populations following treatment policy revisions in the Greater Mekong Subregion"

Wasakul V *et al.*

#### Supplementary material

#### Supplementary Tables

**Supplementary Table 1. Included genetic markers with antimalarial resistance prediction.**

| ***P. falciparum* gene** | **Resistance classification** |
|---|---|
| ***kelch13*[a]**<br>441L, 446I, 449A, 452E, 458Y, 469Y, 469F, 476I, 479I, 481V, 493H, 515K, 522C, 527H, 537I, 537D, 538V, 539T, 543T, 553L, 561H, 568G, 574L, 575K, 579I, 580Y, 584V, 667T, 673I, 675V or 719N as homozygous call | artemisinin |
| ***plasmepsin2/3* (*pm23*) amplification[b]**<br>***WHO kelch13 mutant and multiple copies of pm23*** | piperaquine<br>dihydroartemisinin-piperaquine (DHA-PPQ) |
| ***crt***<br>76T | chloroquine |
| ***dhfr***<br>108N<br>51I and 59R and 108N, all homozygous | <br>pyrimethamine<br>sulfadoxine-pyrimethamine |
| ***dhps***<br>437G | <br>sulfadoxine |
| ***dhfr* & *dhps***<br>*dhfr*: 51I + 59R + 108N + *dhps*: 437G + 540E<br>+ one of *dhfr*:164L, *dhps*:581G, *dhps*:613S or *dhps*:613T<br>with all mutants homozygous | <br>sulfadoxine-pyrimethamine (IPTp[c]) |
| ***mdr1* amplification[b]**<br>***WHO kelch13 mutant and multiple copies of mdr1*** | mefloquine<br>artesunate-mefloquine |

Comprehensive details of genetic markers used in SPOTMalaria: https://ngs.sanger.ac.uk/production/malaria/Resource/29/20200705-GenRe-04a-SpotMalaria-0.39.pdf, pages 4-7, and phenotype prediction rules used in the GenRe-Mekong project: https://ngs.sanger.ac.uk/production/malaria/Resource/29/20200705-GenRe-05-PhenotypeRules-0.39.pdf

[a] The list of validated mutations in the BTB/POZ and the propeller domain was based on the WHO list: https://iris.who.int/handle/10665/274362
[b] Confirmatory *plasmepsin2/3* and *mdr1* amplification testing was performed using qPCR.
[c] Intermittent preventive treatment in pregnancy

**Supplementary Table 2. Genotypically confirmed *P. falciparum* samples by year and province.**

| Country | Province | 2017 | 2018 | 2019 | 2020 | 2021 | 2022 |
|---|---|---|---|---|---|---|---|
| Cambodia | Kampong Speu | | | | 20 | 38 | 8 |
| | Mondulkiri | | | | 4 | 7 | 7 |
| | Pursat | | | | 6 | 19 | 84 |
| | Ratanakiri | 26 | | | | | |
| | Stung Treng | 23 | | | | | |
| Laos | Attapeu | 101 | 208 | 48 | 210 | 122 | 59 |
| | Champasak | 94 | 147 | 28 | 2 | 11 | |
| | Salavan | 103 | 99 | 27 | 15 | | 2 |
| | Savannakhet | 284 | 387 | 212 | 70 | 116 | 8 |
| | Sekong | 13 | 9 | 12 | 9 | 4 | |
| Vietnam | Binh Phuoc | 51 | 324 | 24 | 3 | 4 | |
| | Binh Thuan | | | | 7 | 1 | |
| | Dak Lak | 167 | 125 | 298 | 46 | | 2 |
| | Dak Nong | 60 | 70 | 48 | 7 | | |
| | Gia Lai | 272 | 446 | 523 | 151 | 73 | 144 |
| | Khanh Hoa | 26 | 48 | 7 | 1 | | |
| | Kon Tum | | | 3 | | | |
| | Ninh Thuan | 43 | 12 | 13 | 1 | | 2 |
| | Phu Yen | | | 197 | 55 | 17 | 9 |
| | Quang Binh | | | | | | 1 |
| | Quang Tri | 28 | 19 | 7 | 1 | 1 | 3 |

**Supplementary Table 3. Distribution of *kelch13* allele variants.**

The proportion of samples with kelch13 variant in each country are shown as percentages for each year; all detected variants are shown. "WT" (wild type) indicates no mutation was detected in *kelch13*. "Heterozygous" indicates samples containing multiple parasites genomes carrying more than one *kelch13* mutations. "Missing" indicates parasites whose *kelch13* genotype could not be determined. "Resistant" status was determined from the list of validated alleles associated with delayed clearance, as published by WHO.[1] Full details about the classification method are is given in the SpotMalaria Technical Notes at https://www.malariagen.net/resource/29.

| Country | Classification | *kelch13 allele* | 2017 | 2018 | 2019 | 2020 | 2021 | 2022 |
|---|---|---|---|---|---|---|---|---|
| **Cambodia** | Sensitive | WT | 10.2% | | | 10.0% | 1.6% | 7.1% |
| | | A578S | | | | | | 2.0% |
| | Resistant | C580Y | 83.7% | | | 30.0% | 54.7% | 5.1% |
| | | P553L | 4.1% | | | | | |
| | | Y493H | | | | 36.7% | 29.7% | 67.7% |
| | Undetermined | missing | 2.0% | | | 23.3% | 4.7% | 7.1% |
| | | heterozygous | | | | | 9.4% | 11.1% |
| **Laos** | Sensitive | WT | 43.2% | 64.4% | 30.0% | 24.2% | 42.7% | 13.0% |
| | Resistant | C580Y | 12.9% | 22.5% | 15.6% | 12.7% | 17.4% | 49.3% |
| | | P574L | 0.8% | | | | | |
| | | R539T | 0.2% | 1.2% | 3.4% | 45.4% | 27.3% | 24.6% |
| | | Y493H | 1.5% | 0.1% | | 0.3% | | |
| | Undetermined | missing | 40.5% | 10.0% | 44.6% | 16.0% | 7.9% | 13.0% |
| | | heterozygous | 0.8% | 1.9% | 6.4% | 1.3% | 4.0% | |
| | | G357S | | | | | 0.4% | |
| | | G544R | | | | | 0.4% | |
| **Vietnam** | Sensitive | WT | 20.2% | 11.8% | 5.0% | 0.7% | 2.1% | 2.5% |
| | Resistant | C469F | 0.5% | 0.1% | | | | |
| | | C580Y | 54.4% | 77.3% | 86.0% | 89.3% | 95.8% | 77.6% |
| | | P553L | 1.5% | 0.3% | | | | |
| | | R539T | 0.2% | | | | | |
| | Undetermined | missing | 20.6% | 8.6% | 8.0% | 9.6% | 2.1% | 18.6% |
| | | heterozygous | 2.6% | 1.9% | 1.0% | 0.4% | | 1.2% |

**Supplementary Table 4. Changes in PPQ-R proportions in the three largest KEL1/PLA1 clusters.**
The proportion of samples predicted to be resistant to piperaquine (PPQ) in the largest three KEL1/PLA1 clusters (KLV01, KLV02, KLV03) are shown, aggregated by quarter (Q1: January-March; Q2: April-June; Q3: July-September; Q4: October-December). "NA" means there are no samples from the cluster in a given period.

| | | PPQ-R | | |
|---|---|---|---|---|
| | | **KLV01** | **KLV02** | **KLV03** |
| 2017 | Q1 | **100%** | **NA** | **NA** |
| | Q2 | **100%** | **NA** | **NA** |
| | Q3 | **86%** | **100%** | **NA** |
| | Q4 | **86%** | **100%** | **NA** |
| 2018 | Q1 | **96%** | **100%** | **NA** |
| | Q2 | **94%** | **100%** | **NA** |
| | Q3 | **100%** | **98%** | **100%** |
| | Q4 | **96%** | **100%** | **100%** |
| 2019 | Q1 | **92%** | **100%** | **100%** |
| | Q2 | **96%** | **98%** | **90%** |
| | Q3 | **99%** | **91%** | **91%** |
| | Q4 | **96%** | **98%** | **87%** |
| 2020 | Q1 | **94%** | **95%** | **93%** |
| | Q2 | **82%** | **100%** | **85%** |
| | Q3 | **20%** | **100%** | **67%** |
| | Q4 | **17%** | **NA** | **30%** |
| 2021 | Q1 | **0%** | **NA** | **25%** |
| | Q2 | **0%** | **NA** | **8%** |
| | Q3 | **NA** | **NA** | **0%** |
| | Q4 | **NA** | **NA** | **0%** |
| 2022 | Q1 | **NA** | **NA** | **0%** |
| | Q2 | **NA** | **NA** | **0%** |
| | Q3 | **NA** | **NA** | **0%** |
| | Q4 | **NA** | **NA** | **0%** |

**Supplementary Table 5. Temporal cluster distribution of *P. falciparum* samples.**

For each cluster, we show: the cluster label; proportion of samples per year of collection; number of sample in a cluster, proportion of samples predicted resistant to artemisinin (ART), piperaquine (PPQ), and mefloquine (MQ); chloroquine (CQ), sulfadoxine (SX) and pyrimethamine (PM); number of sample with genotyped kelch13 variant; whether the cluster is a KEL1/PLA1 haplotype. All clusters of at least 10 members are shown. Samples that could not be assigned to a cluster with these parameters were labelled as 'not clustered'.

| | Proportion of samples in each year | | | | | | | Resistance | | | | | | | |
|---|---|---|---|---|---|---|---|---|---|---|---|---|---|---|---|
| **Cluster** | **2017** | **2018** | **2019** | **2020** | **2021** | **2022** | **Sample Count** | **ART** | **PPQ** | **MQ** | **CQ** | **SX** | **PM** | **Kelch13** | **KEL1/PLA1** |
| KLV001 | 5% | 8% | 21% | 16% | 4% | | 635 | 100% | 89% | 0% | 100% | 100% | 100% | C580Y:622 | Yes |
| KLV002 | 1% | 12% | 18% | 4% | | | 528 | 100% | 98% | 0% | 100% | 100% | 100% | C580Y:524 | Yes |
| KLV003 | | 1% | 13% | 10% | 16% | 33% | 450 | 100% | 54% | 0% | 100% | 100% | 100% | C580Y:449 | Yes |
| KLV004 | 0% | 0% | 0% | 18% | 15% | 4% | 202 | 100% | 0% | 13% | 100% | 100% | 100% | R539T:190; C580Y:1 | |
| KLV005 | 4% | 4% | 3% | 3% | 0% | | 179 | 100% | 78% | 0% | 100% | 100% | 100% | C580Y:157 | Yes |
| KLV006 | 5% | 3% | 1% | 1% | | | 153 | 6% | 1% | 0% | 100% | 0% | 100% | WT:116; Y493H:6; C580Y:1; P574L:1 | |
| KLV007 | 1% | 1% | 0% | 3% | 10% | 9% | 114 | 100% | 1% | 0% | 100% | 100% | 100% | C580Y:108 | |
| KLV008 | 5% | 2% | 0% | | | | 107 | 100% | 84% | 0% | 100% | 100% | 100% | C580Y:92 | Yes |
| KLV009 | 1% | 1% | 3% | 4% | | | 88 | 100% | 100% | 0% | 100% | 100% | 100% | C580Y:86 | Yes |
| KLV010 | 3% | 2% | 0% | 0% | | | 83 | 1% | 0% | 0% | 100% | 0% | 100% | WT:68; C580Y:1 | |
| KLV011 | | | 0% | | 14% | | 60 | 0% | 0% | 0% | 0% | 0% | 100% | WT:59 | |
| KLV012 | | | | | 1% | 16% | 56 | 100% | 0% | 0% | 100% | 100% | 100% | Y493H:56 | |
| KLV013 | 2% | 1% | 1% | | | | 53 | 0% | 2% | 0% | 100% | 100% | 100% | WT:50 | |
| KLV014 | 0% | 1% | 2% | 1% | | | 47 | 100% | 85% | 0% | 100% | 100% | 100% | C580Y:46 | Yes |
| KLV015 | | 2% | 0% | 0% | | | 44 | 0% | 0% | 0% | 100% | 100% | 100% | WT:43 | |
| KLV016 | 1% | 1% | | | 0% | | 41 | 8% | 0% | 0% | 100% | 0% | 100% | WT:36; C580Y:2; P574L:1 | |
| KLV017 | | 1% | 0% | 3% | | | 37 | 0% | 0% | 0% | 100% | 0% | 100% | WT:36 | |
| KLV018 | 2% | 1% | 0% | | | | 35 | 100% | 97% | 0% | 100% | 100% | 100% | C580Y:28 | Yes |
| KLV019 | 0% | 1% | 0% | | | | 28 | 0% | 0% | 0% | 100% | 0% | 100% | WT:19 | |
| KLV020 | | 1% | | 0% | | | 27 | 0% | 0% | 0% | 100% | 0% | 100% | WT:27 | |
| KLV021 | | 1% | 0% | | | | 26 | 0% | 0% | 0% | 100% | 100% | 100% | WT:25 | |
| KLV022 | 1% | 0% | 1% | | | | 26 | 0% | 0% | 0% | 100% | 100% | 100% | WT:23 | |
| KLV023 | | | | 1% | 4% | 1% | 25 | 100% | 0% | 0% | 100% | 100% | 100% | C580Y:25 | |
| KLV024 | 1% | 1% | | | | | 22 | 100% | 100% | 0% | 100% | 100% | 100% | C580Y:19 | Yes |
| KLV025 | 1% | 1% | 0% | | | | 22 | 75% | 0% | 0% | 100% | 100% | 100% | C580Y:12; WT:4 | |
| KLV026 | 0% | | 1% | 1% | | | 20 | 100% | 94% | 0% | 100% | 100% | 100% | C580Y:20 | Yes |
| KLV027 | 1% | 0% | 0% | | | | 20 | 0% | 7% | 0% | 100% | 100% | 100% | WT:17 | |
| KLV028 | | 0% | 0% | 1% | | | 19 | 0% | 0% | 0% | 100% | 100% | 100% | WT:19 | |
| KLV029 | 1% | 0% | 0% | | | | 19 | 16% | 0% | 0% | 0% | 100% | 100% | WT:16; C580Y:3 | |
| KLV030 | 0% | 1% | 0% | | | | 19 | 100% | 0% | 0% | 100% | 100% | 100% | C580Y:15 | |
| KLV031 | | | 0% | 2% | 0% | | 18 | 0% | 0% | 0% | 100% | 0% | 0% | WT:16 | |
| KLV032 | | 0% | 0% | 0% | 3% | | 18 | 0% | 0% | 0% | 0% | 0% | 100% | WT:17 | |
| KLV033 | | 1% | 0% | 0% | 0% | | 18 | 0% | 0% | 0% | 100% | 0% | 100% | WT:17 | |
| KLV034 | 1% | 1% | | | | | 18 | 0% | 7% | 0% | 100% | 100% | 100% | WT:16 | |
| KLV035 | 1% | 0% | | | | | 18 | 12% | 0% | 0% | 100% | 100% | 100% | WT:7; C580Y:1 | |
| KLV036 | | | | | 3% | 1% | 17 | 0% | 0% | 0% | 100% | 0% | 100% | WT:17 | |
| KLV037 | 1% | 0% | 0% | | | | 17 | 8% | 0% | 27% | 0% | 100% | 100% | WT:11; C580Y:1 | |
| KLV038 | | | 0% | 1% | 0% | | 16 | 100% | 0% | 0% | 100% | 100% | 100% | R539T:15 | |
| KLV039 | | 1% | 0% | | | | 16 | 100% | 42% | 0% | 100% | 100% | 100% | C580Y:16 | |
| KLV040 | 0% | | 1% | 0% | | | 16 | 100% | 0% | 0% | 100% | 100% | 100% | C580Y:14 | |
| KLV041 | 1% | 0% | 0% | | | | 16 | 0% | 0% | 0% | 100% | 0% | 100% | WT:9 | |
| KLV042 | 0% | 1% | | | | | 15 | 100% | 0% | 0% | 100% | 100% | 100% | C580Y:14 | |
| KLV043 | | | | 2% | 0% | 1% | 13 | 100% | 0% | 0% | 100% | 100% | 100% | Y493H:13 | |
| KLV044 | 0% | 1% | | | | | 13 | 0% | 0% | 0% | 100% | 0% | 100% | WT:12 | |
| KLV045 | | 1% | | | | | 13 | 0% | 0% | 0% | 100% | 0% | 100% | WT:13 | |
| KLV046 | 1% | 0% | | | | | 13 | 55% | 0% | 0% | 100% | 0% | 100% | C580Y:6; WT:5 | |
| KLV047 | | 1% | | | | | 12 | 0% | 0% | 0% | 100% | 0% | 100% | WT:12 | |
| KLV048 | | 1% | | | | | 12 | 0% | 20% | 0% | 100% | 0% | 100% | WT:12 | |
| KLV049 | 0% | 0% | 0% | | | | 12 | 100% | 100% | 0% | 100% | 100% | 100% | C580Y:12 | Yes |
| KLV050 | 0% | 0% | | 0% | | | 12 | 100% | 0% | 0% | 100% | 100% | 100% | C580Y:10 | |
| KLV051 | 0% | 0% | 0% | 1% | | | 12 | 0% | 0% | 0% | 0% | 0% | 100% | WT:9 | |
| KLV052 | 1% | 0% | | | | | 12 | 100% | 100% | NA | 100% | 100% | 100% | C580Y:5 | Yes |
| KLV053 | 1% | 0% | | | | | 12 | 33% | 11% | 0% | 100% | 100% | 100% | WT:2; C580Y:1 | |
| KLV054 | 1% | 0% | | | | | 12 | 100% | 0% | 0% | 100% | 100% | 100% | P553L:11 | |
| KLV055 | | 1% | | | | | 11 | 0% | 0% | 0% | 0% | 0% | 0% | WT:11 | |
| KLV056 | 1% | 0% | | 0% | | | 11 | 22% | 0% | 0% | 100% | 100% | 100% | WT:7; Y493H:2 | |
| KLV057 | 0% | 0% | 0% | 1% | | | 11 | 0% | 0% | 0% | 100% | 0% | 100% | WT:9 | |
| KLV058 | 0% | 0% | | | | | 11 | 100% | 100% | 0% | 100% | 100% | 100% | C580Y:11 | Yes |
| KLV059 | 1% | | | | | | 11 | 0% | 0% | NA | 0% | 100% | 100% | WT:5 | |
| KLV060 | | 1% | | | | | 10 | 0% | 0% | NA | 100% | 0% | 100% | WT:10 | |
| KLV061 | | 1% | | | | | 10 | 100% | 0% | 0% | 100% | 100% | 100% | C580Y:10 | |
| KLV062 | | 1% | | | | | 10 | 0% | 0% | 0% | 100% | 0% | 100% | WT:7 | |
| KLV063 | 0% | 0% | 0% | | | | 10 | 10% | 0% | 0% | 100% | 0% | 100% | WT:9; P574L:1 | |
| KLV064 | 1% | | | | | | 10 | 0% | 0% | NA | 0% | 0% | 100% | WT:10 | |
| Not clustered | 55% | 43% | 30% | 26% | 26% | 36% | | | | | | | | | |
| Total samples | 1291 | 1894 | 1447 | 608 | 413 | 329 | | | | | | | | | |

#### Supplementary Figures

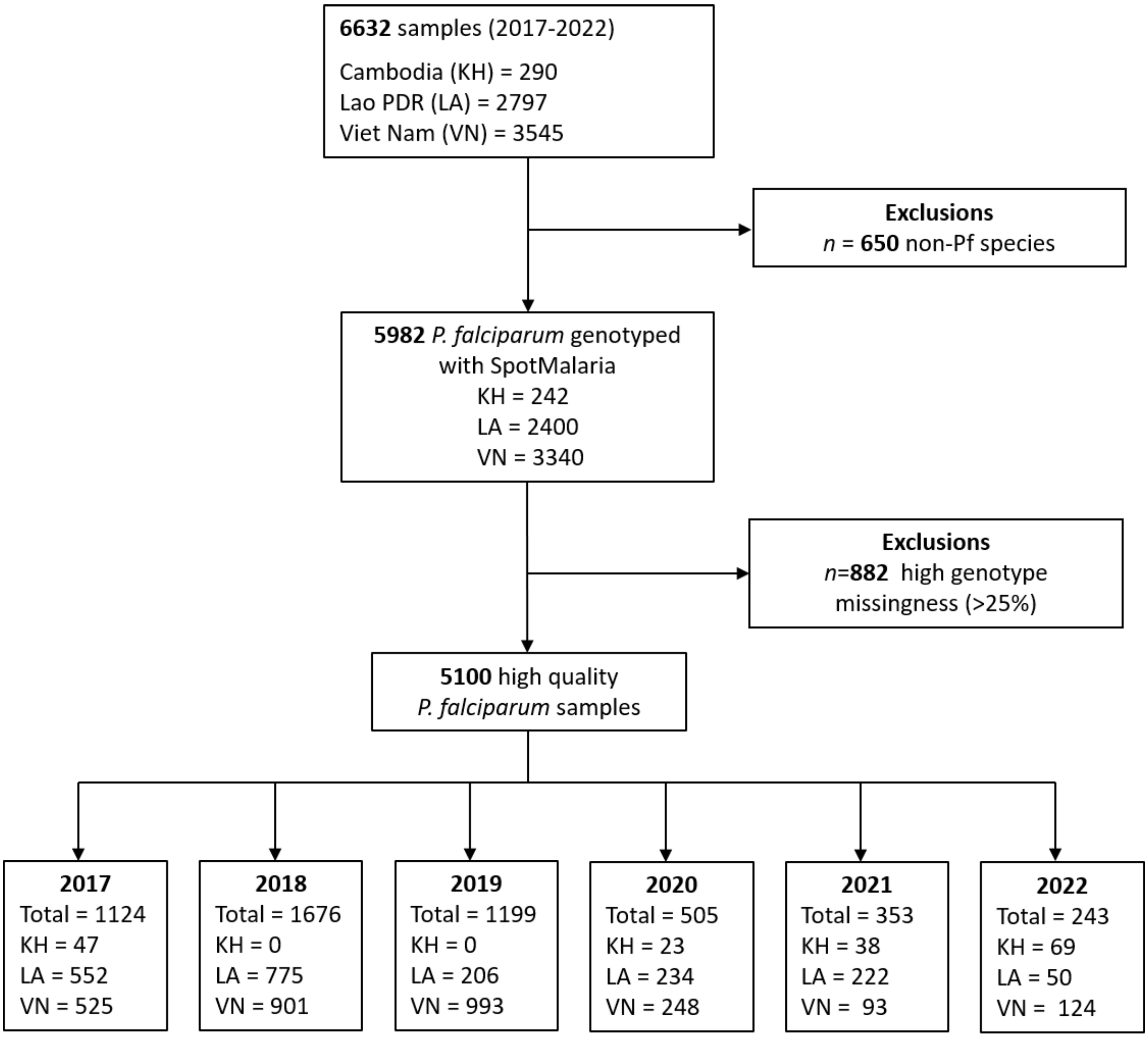

**Supplementary Figure 1. Collected and genotyped *P. falciparum* (*Pf*) samples.**
The diagram shows how the analysed sample set was derived from *Pf* samples collected by GenRe-Mekong, and the composition of this sample set by country and year.

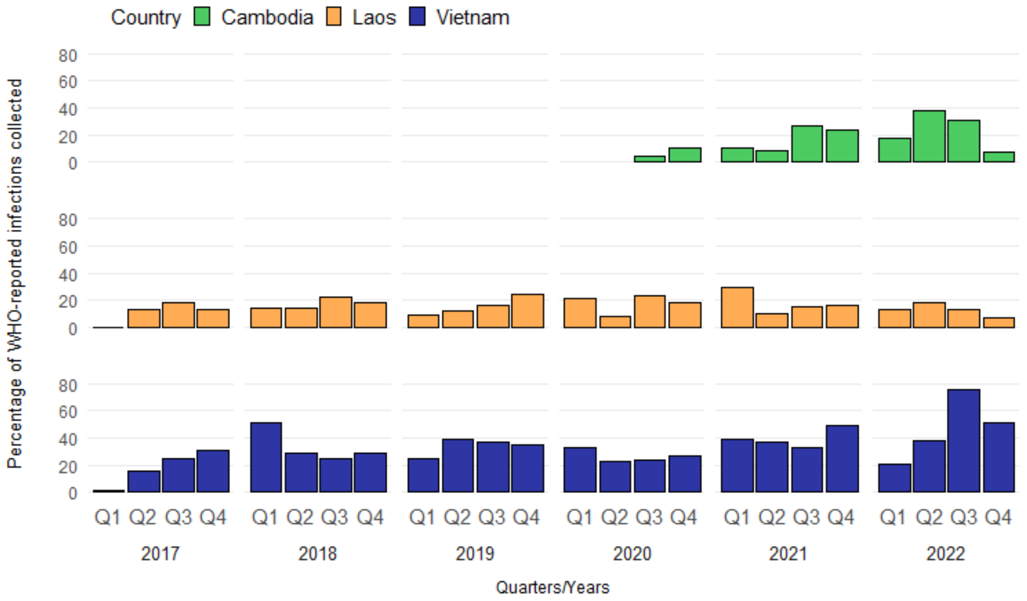

**Supplementary Figure 2. Percentage of WHO-registered *Pf* infections processed by GenRe-Mekong.** Between January 2017 to December 2022, 32.7% and 16.2% WHO-reported *Pf* infections in Vietnam and Laos were sampled by the GenRe-Mekong project, respectively. For Cambodia, 15.6% of WHO-reported *P. falciparum* infections occurred between 2020 and 2022 were sampled GenRe-Mekong.[2-4]

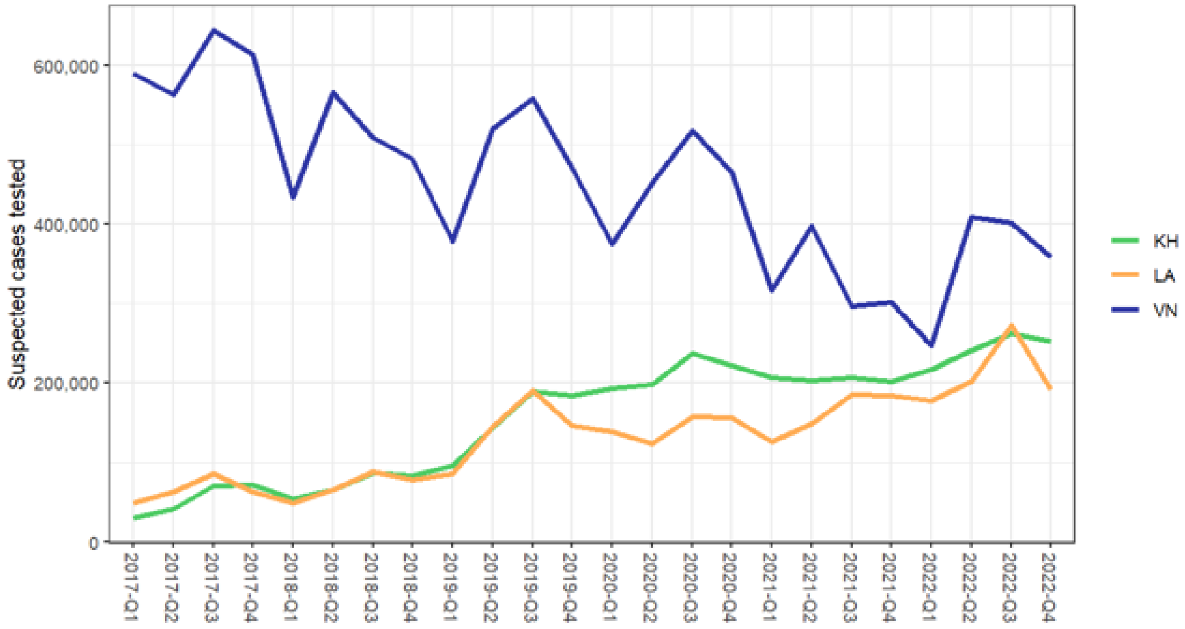

**Supplementary Figure 3. Number of suspected cases tested in each country.**
Numbers reported to the WHO Mekong Malaria Elimination Programme.[2-4] KH: Cambodia, LA: Lao PDR and VN: Vietnam.

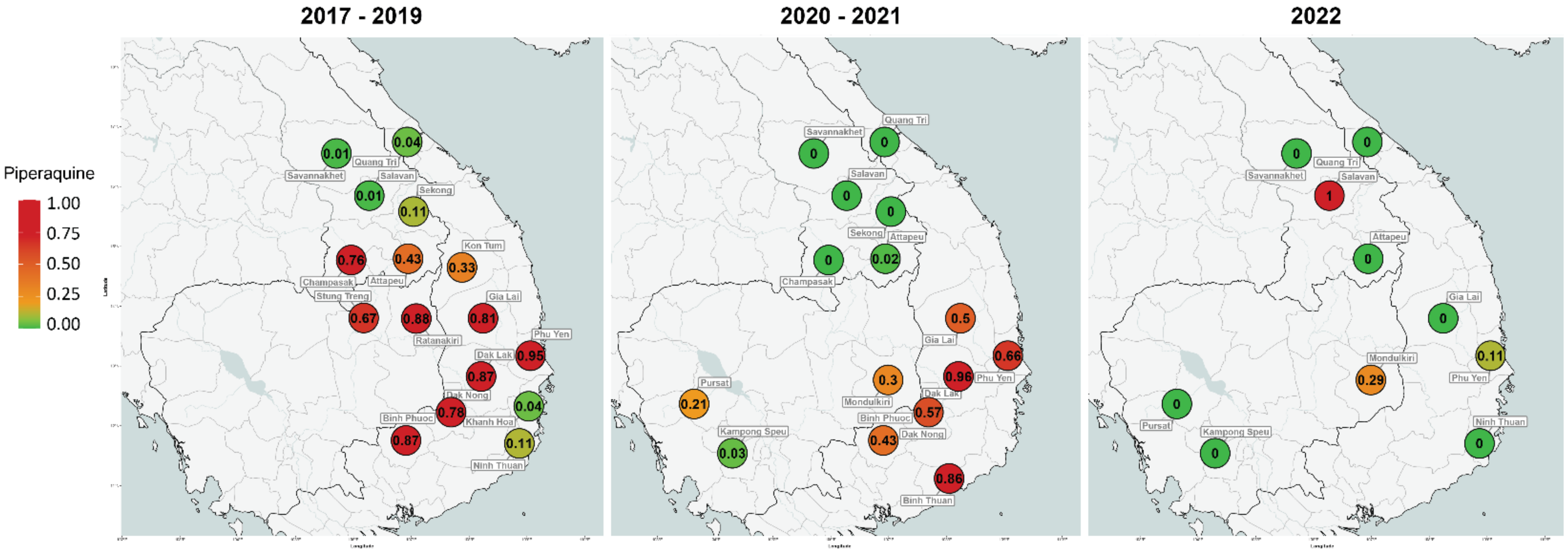

**Supplementary Figure 4. Frequency of piperaquine-resistant parasites over three time periods.**

The predictor marker for piperaquine resistance was the *pm23* amplifications. Left panel: 2017-2019, middle: 2020-2021, right: January-December 2022. Marker colours reflect resistance prevalence, ranging from 0 to 1, where 0 means no parasite were predicted to be resistant, and 1 means 100% of the parasites in the province carried the relevant resistance markers. A marker is shown only if there are at least 2 samples from the province (e.g.: only two samples were samples were present in Salavan in 2022).

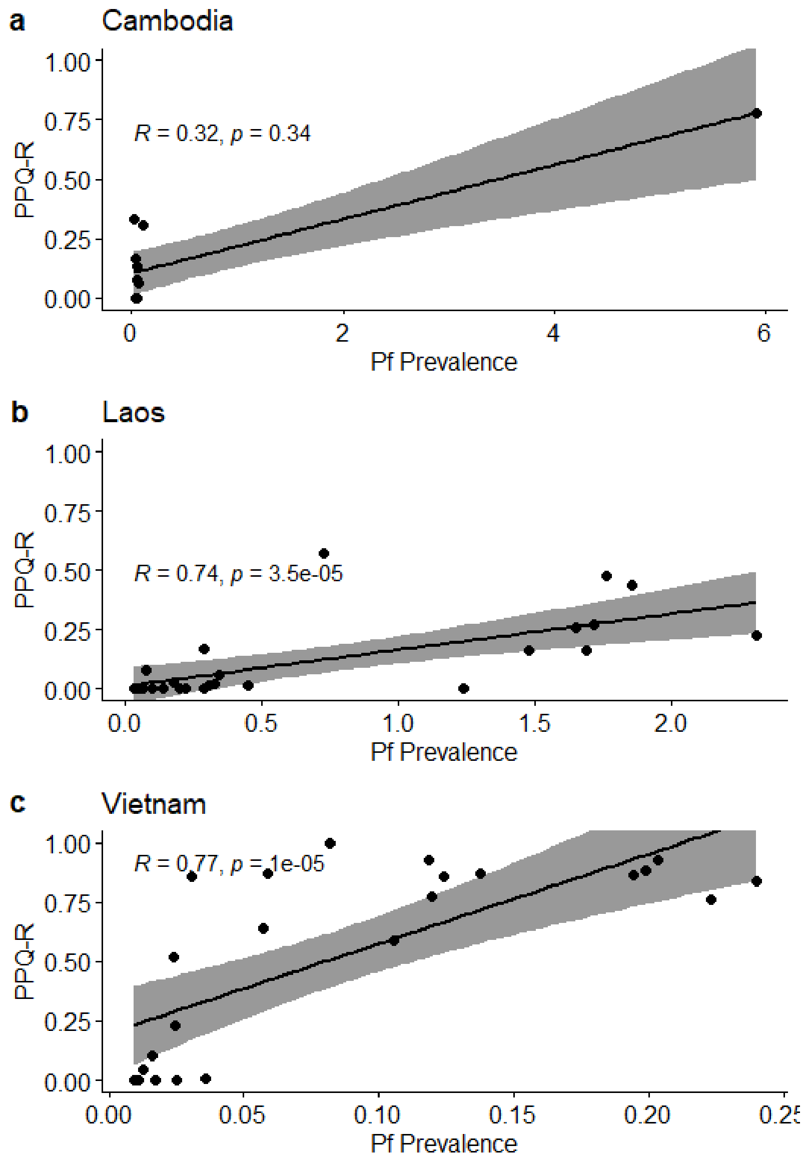

**Supplementary Figure 5. Relationship between PPQ resistance (PPQ-R) and *Pf* prevalence.**
For each country, we show a Spearman's rank correlation analysis highlighting the relationship between levels of PPQ resistance (PPQ-R) and prevalence of *Plasmodium falciparum* (*Pf*) across three countries: (a) Cambodia, (b) Laos, and (c) Vietnam. Data points represents quarterly observations (n = 24 per country) over six years. Each panel shows the linear regression fit (black line) with 95% confidence intervals (shaded gray area). The correlation coefficient (R) and associated p-value are shown in each panel. The analysis indicates a weak non-significant correlation in Cambodia, but strong and significant positive correlations in Laos and Vietnam. *Pf* malaria prevalence was estimated from WHO data.2-4

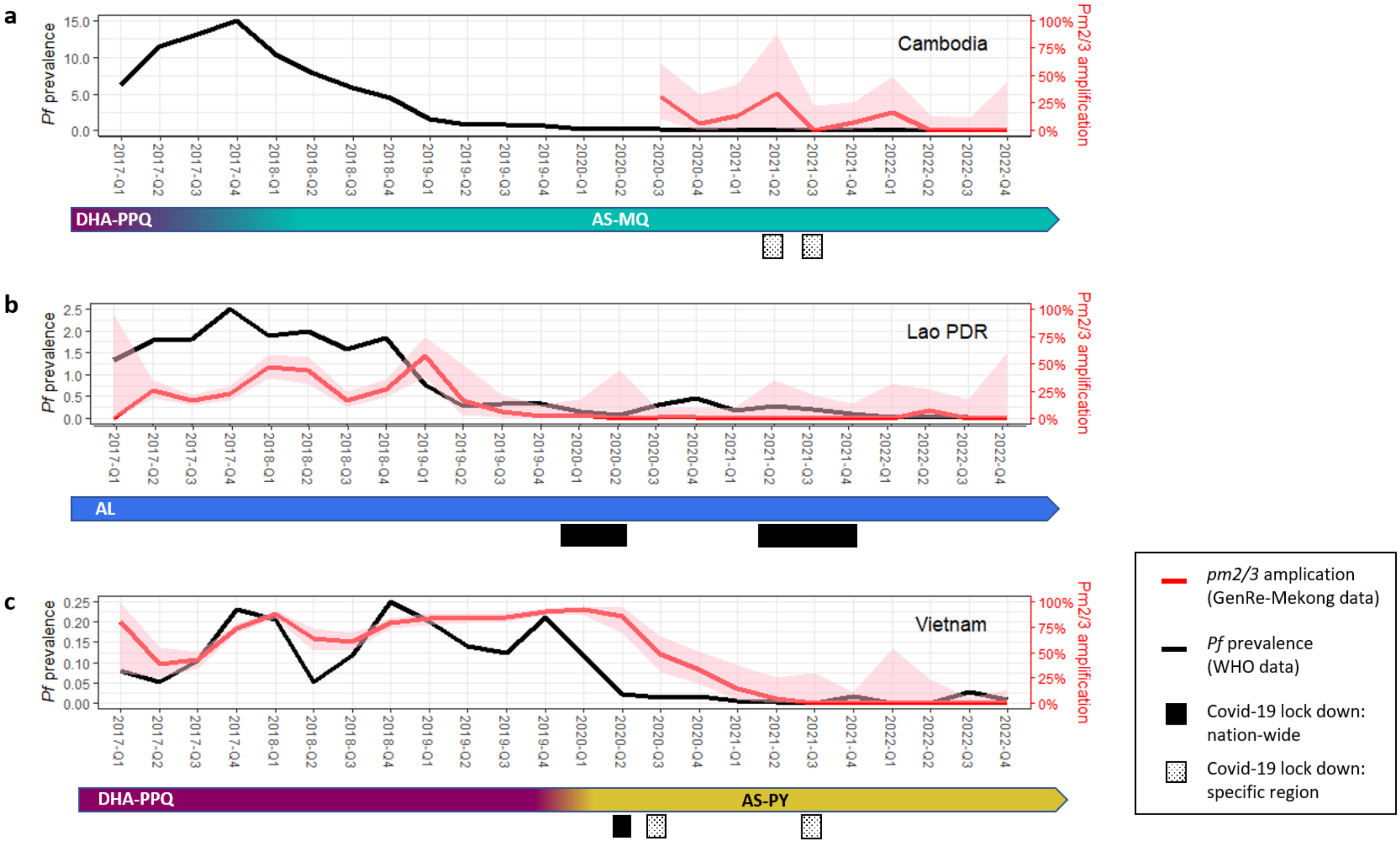

**Supplementary Figure 6. Prevalence and *pm2/3* amplifications frequency changes.**
Panels: (**a**) Cambodia, (**b**) Laos and (**c**) Vietnam. *Pf* malaria prevalence (black lines, left-axis) was derived from WHO data,[2-4] while *pm23* amplification frequencies (red lines, right-axis) were derived from GenRe-Mekong genotypes. Shaded areas represent 95% confidence intervals, with wider intervals reflecting smaller sample sizes. The *Pf* prevalence scale varies between countries. First line ACT treatment for uncomplicated malaria, according to national policy, and COVID-19 lockdown restrictions are shown under each graph. Cambodia switched ACT from dihydroartemisinin-piperaquine (DHA-PPQ) to artesunate-mefloquine (AS-MQ) starting 2017,[5,6] and did not implement a nationwide lockdown but focused on localized restrictions: in Phnom Penh and Ta Khmau in April-May 2021, and provinces bordering Thailand in July-August 2021.[7] GenRe-Mekong started routine surveillance in Cambodia in Q3 2020. Laos continued the use of artemether-lumefantrine (AL) throughout the study period, and lockdowns were imposed nationally March-May 2020[8] and April-December 2021.[9] Vietnam changed frontline ACT from DHA-PPQ to artesunate-pyronaridine (AS-PYR) in five provinces in late Q1 2020,[10,11] and COVID-19 lockdowns were imposed nationwide in April-May 2020; in Danang July-August 2020; and in Hanoi and the Southern region in July-August 2021.[12,13]

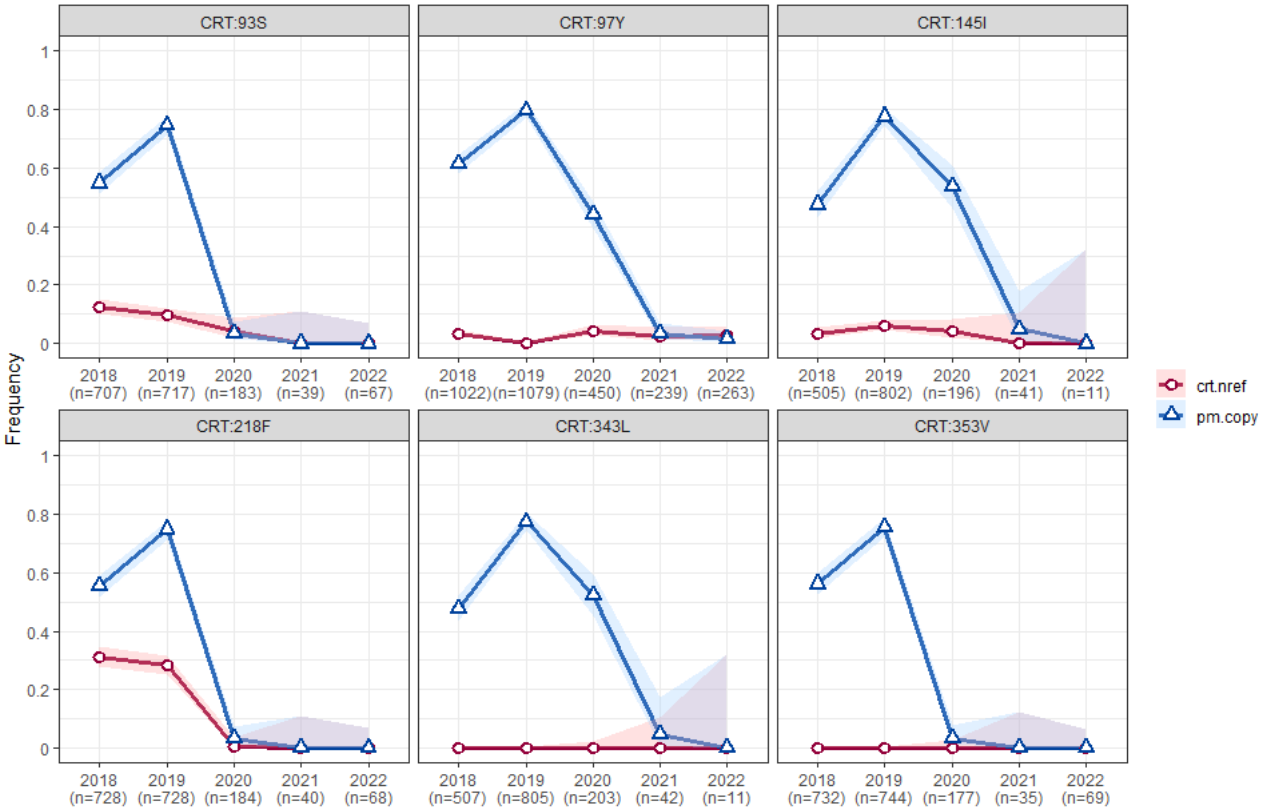

**Supplementary Figure 7. Temporal distribution of *crt* mutations and *pm23* amplifications**.
The proportion of samples carrying *crt* mutations is shown by year in red (crt.nref), while the proportion of *pm23* amplifications is shown in blue (pm.copy), with value ranging from 0 (no samples carrying the resistant allele) to 1 (100% of samples carrying the resistant allele). Sample sizes (n) are indicated on the x-axis for each year. Shaded areas represent 95% confidence intervals, with wider intervals reflecting smaller sample sizes. Analysis includes only samples for which both the *crt* and *pm23* genotypes are available.

**(Next Page)**

**Supplementary Figure 8. Temporal distribution of crt mutations and pm23 amplifications.**
The proportion of samples carrying *crt* mutations is shown by year in red (crt.nref), while the proportion of *pm23* amplifications is shown in blue (pm.copy) for (**a**) Cambodia (KH), (**b**) Laos (LA) and (**c**) Vietnam (VN), with value ranging from 0 (no samples carrying the resistant allele) to 1 (100% of samples carrying the resistant allele). Sample sizes (n) are indicated on the x-axis for each year. Shaded areas represent 95% confidence intervals, with wider intervals reflecting smaller sample sizes. Analysis includes only samples for which both the *crt* and *pm23* genotypes are available.

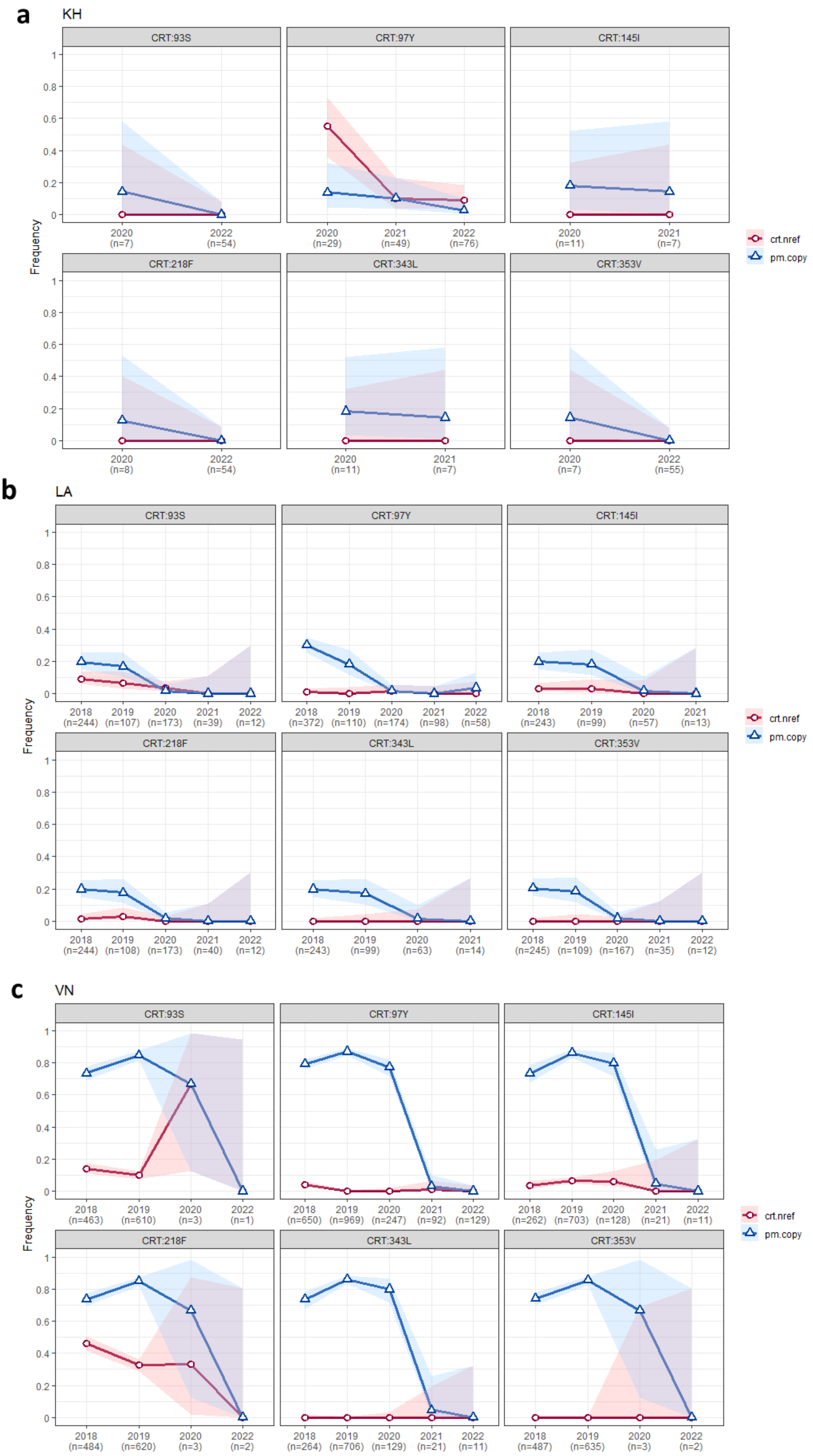
a
KH
b
LA
c
VN
CRT:93S
CRT:97Y
CRT:145I
CRT:218F
CRT:343L
CRT:353V
Frequency
crt.nref
pm.copy
2020 (n=7)
2022 (n=54)
2020 (n=29)
2021 (n=49)
2022 (n=76)
2020 (n=11)
2021 (n=7)
2020 (n=8)
2022 (n=54)
2020 (n=11)
2021 (n=7)
2020 (n=7)
2022 (n=55)
2018 (n=244)
2019 (n=107)
2020 (n=173)
2021 (n=39)
2022 (n=12)
2018 (n=372)
2019 (n=110)
2020 (n=174)
2021 (n=98)
2022 (n=58)
2018 (n=243)
2019 (n=99)
2020 (n=57)
2021 (n=13)
2018 (n=244)
2019 (n=108)
2020 (n=173)
2021 (n=40)
2022 (n=12)
2018 (n=243)
2019 (n=99)
2020 (n=63)
2021 (n=14)
2018 (n=245)
2019 (n=109)
2020 (n=167)
2021 (n=35)
2022 (n=12)
2018 (n=463)
2019 (n=610)
2020 (n=3)
2022 (n=1)
2018 (n=650)
2019 (n=969)
2020 (n=247)
2021 (n=92)
2022 (n=129)
2018 (n=262)
2019 (n=703)
2020 (n=128)
2021 (n=21)
2022 (n=11)
2018 (n=484)
2019 (n=620)
2020 (n=3)
2022 (n=2)
2018 (n=264)
2019 (n=706)
2020 (n=129)
2021 (n=21)
2022 (n=11)
2018 (n=487)
2019 (n=635)
2020 (n=3)
2022 (n=2)

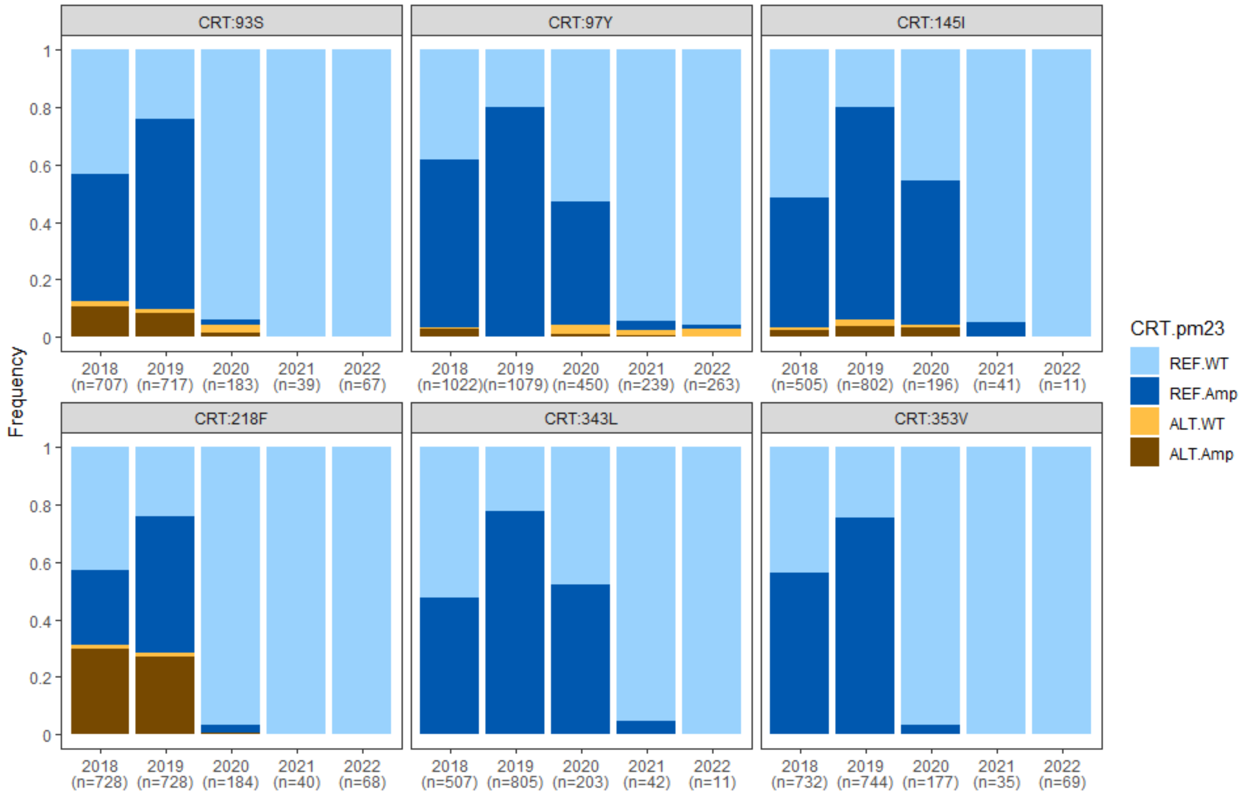

**Supplementary Figure 9. Regional proportions of samples with *crt* mutations and *pm23* amplifications.**
The stacked bar plots show the proportion of each genotype combination in each year: wild-type for both *crt* and *pm23* (REF.WT, light blue), wild-type *crt* with *pm23* amplification (REF.Amp, navy)**,** *crt* mutant with single-copy *pm23* (ALT.WT, yellow), and *crt* mutant with *pm23* amplification (ALT.Amp, brown). Sample sizes (n) for each year are shown in parentheses below the x-axis.

**(Next page)**

**Supplementary Figure 10. Proportion of samples with combined *crt* and *pm23* genotypes, by country.**
The stacked bar plots show the proportion of each genotype combination in each year for (**a**) Cambodia (KH), (**b**) Laos (LA) and (**c**) Vietnam (VN): wild-type for both *crt* and *pm23* (REF.WT, light blue), wild-type *crt* with *pm23* amplification (REF.Amp, navy)**,** *crt* mutant with single-copy *pm23* (ALT.WT, yellow), and *crt* mutant with *pm23* amplification (ALT.Amp, brown). Sample sizes (n) for each year are shown in parentheses below the x-axis.

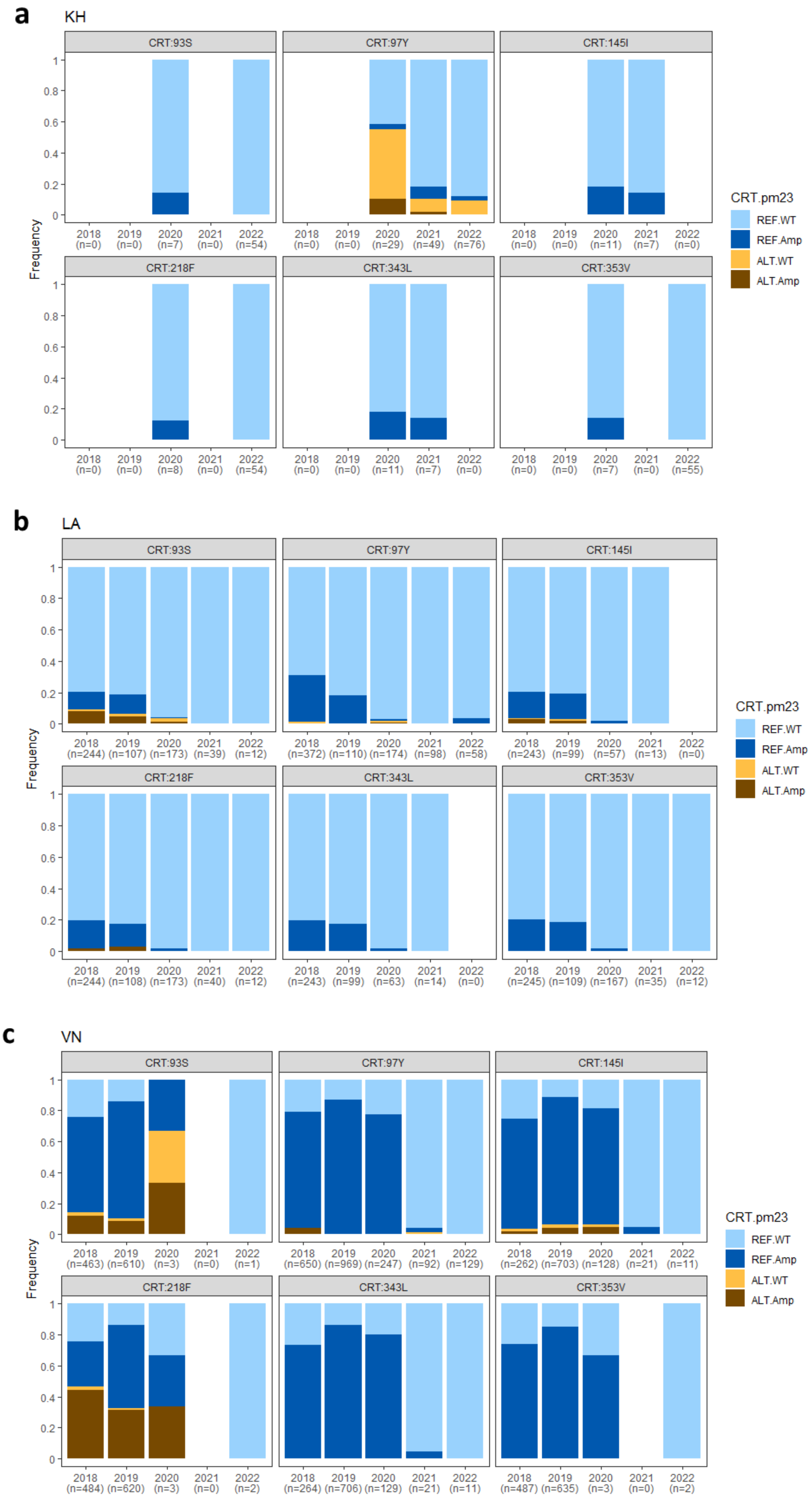

a
KH
CRT:93S
CRT:97Y
CRT:145I
CRT:218F
CRT:343L
CRT:353V
Frequency
2018 (n=0) 2019 (n=0) 2020 (n=7) 2021 (n=0) 2022 (n=54)
2018 (n=0) 2019 (n=0) 2020 (n=29) 2021 (n=49) 2022 (n=76)
2018 (n=0) 2019 (n=0) 2020 (n=11) 2021 (n=7) 2022 (n=0)
2018 (n=0) 2019 (n=0) 2020 (n=8) 2021 (n=0) 2022 (n=54)
2018 (n=0) 2019 (n=0) 2020 (n=11) 2021 (n=7) 2022 (n=0)
2018 (n=0) 2019 (n=0) 2020 (n=7) 2021 (n=0) 2022 (n=55)
CRT.pm23
REF.WT
REF.Amp
ALT.WT
ALT.Amp
b
LA
CRT:93S
CRT:97Y
CRT:145I
CRT:218F
CRT:343L
CRT:353V
Frequency
2018 (n=244) 2019 (n=107) 2020 (n=173) 2021 (n=39) 2022 (n=12)
2018 (n=372) 2019 (n=110) 2020 (n=174) 2021 (n=98) 2022 (n=58)
2018 (n=243) 2019 (n=99) 2020 (n=57) 2021 (n=13) 2022 (n=0)
2018 (n=244) 2019 (n=108) 2020 (n=173) 2021 (n=40) 2022 (n=12)
2018 (n=243) 2019 (n=99) 2020 (n=63) 2021 (n=14) 2022 (n=0)
2018 (n=245) 2019 (n=109) 2020 (n=167) 2021 (n=35) 2022 (n=12)
CRT.pm23
REF.WT
REF.Amp
ALT.WT
ALT.Amp
c
VN
CRT:93S
CRT:97Y
CRT:145I
CRT:218F
CRT:343L
CRT:353V
Frequency
2018 (n=463) 2019 (n=610) 2020 (n=3) 2021 (n=0) 2022 (n=1)
2018 (n=650) 2019 (n=969) 2020 (n=247) 2021 (n=92) 2022 (n=129)
2018 (n=262) 2019 (n=703) 2020 (n=128) 2021 (n=21) 2022 (n=11)
2018 (n=484) 2019 (n=620) 2020 (n=3) 2021 (n=0) 2022 (n=2)
2018 (n=264) 2019 (n=706) 2020 (n=129) 2021 (n=21) 2022 (n=11)
2018 (n=487) 2019 (n=635) 2020 (n=3) 2021 (n=0) 2022 (n=2)
CRT.pm23
REF.WT
REF.Amp
ALT.WT
ALT.Amp

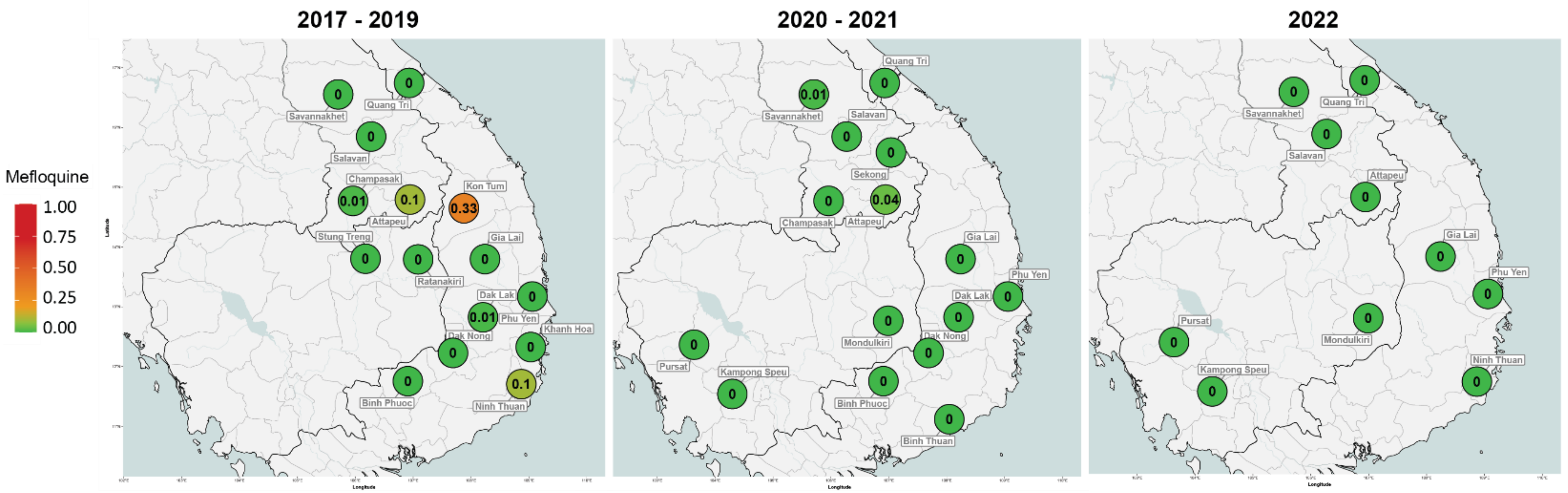

**Supplementary Figure 11. Predicted resistance to mefloquine per province divided in three periods.**
Left panel: 2017-2019, middle: 2020-2021, right: January-December 2022. Marker colours reflects resistance prevalence, ranging from 0 to 1, where 0 means no parasite were predicted to be resistant, and 1 means 100% of the parasites in the province carried the relevant resistance marker. A marker appears when at least 2 samples were processed from the province.

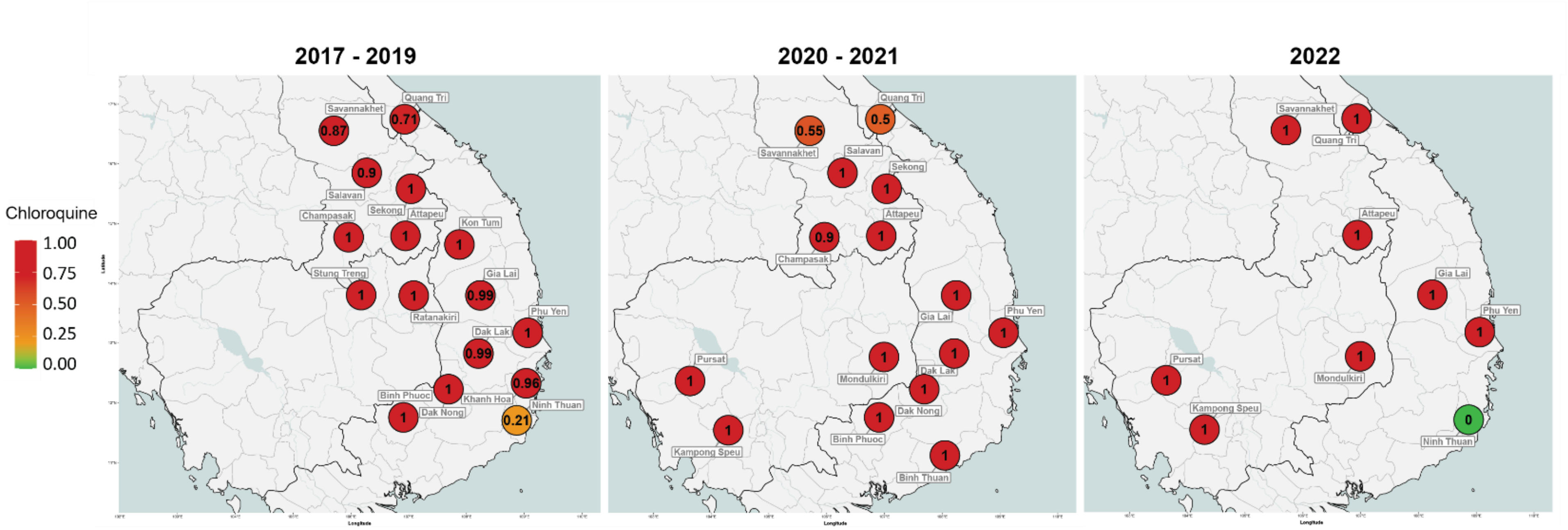

**Supplementary Figure 12. Predicted resistance to chloroquine per province divided in three periods.**

Left panel: 2017-2019, middle: 2020-2021, right: January-December 2022. Marker colours reflects resistance prevalence, ranging from 0 to 1, where 0 means no parasite were predicted to be resistant, and 1 means 100% of the parasites in the province carried the relevant resistance marker. A marker appears when at least 2 samples were processed from the province.

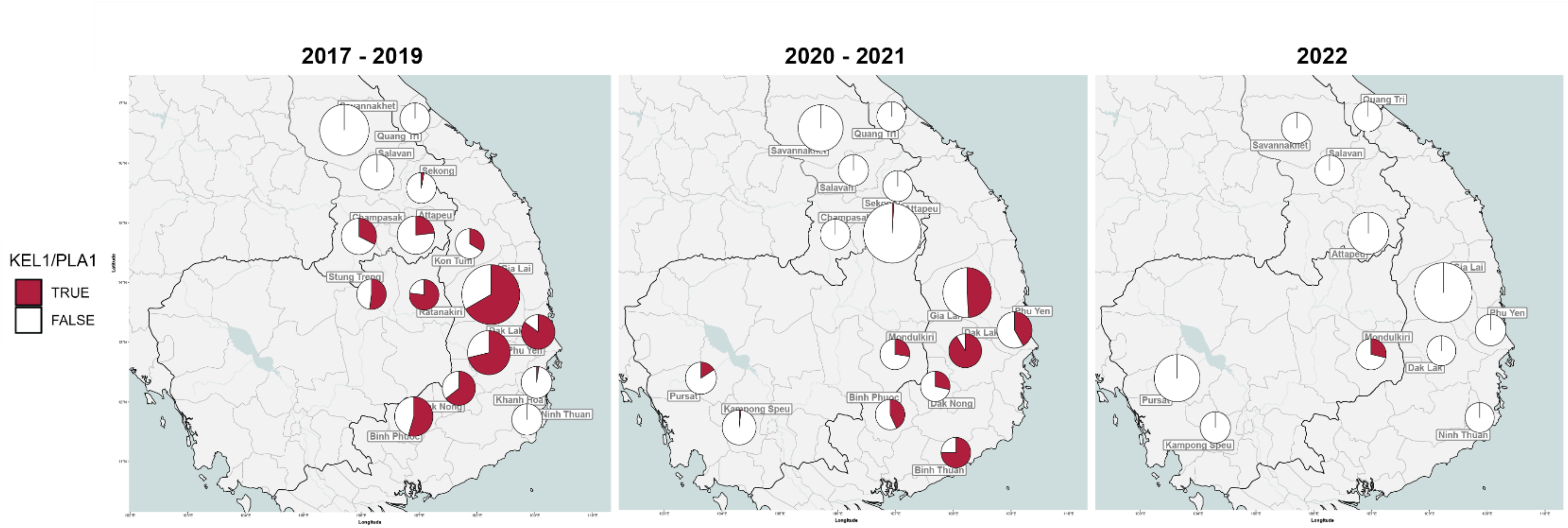

**Supplementary Figure 13. Prevalence of KEL1/PLA1 between 2017 and 2022.**
Pie charts show the proportions of samples classified as KEL1/PLA1, having both C580Y *kelch13* mutation and *plasmepsin 2/3* amplification, in the three periods. Left panel: 2017-2019, middle: 2020-2021, right: January-December 2022. The pie size reflects the sample number in the province.

**(Next page)**

**Supplementary Figure 14. Distribution of *mdr1* haplotypes and *kelch13* alleles.**
Panels show the proportions of parasites carrying different *mdr1* haplotypes (a) and *kelch13* alleles (b) in three time periods (left: 2017-2019, middle: 2020-2021, right: Jan-Dec 2022. NYD is the wild-type *mdr1* haplotype. Parasites with wild-type (WT) *kelch13* alleles are predicted to be sensitive to artemisinin, while other *kelch13* alleles have been associated with delayed parasite clearance and therefore predicted to be artemisinin-resistant, with the exception of G357S and G544R which are not in the WHO validated marker list, and thus their association with artemisinin resistance is undetermined.

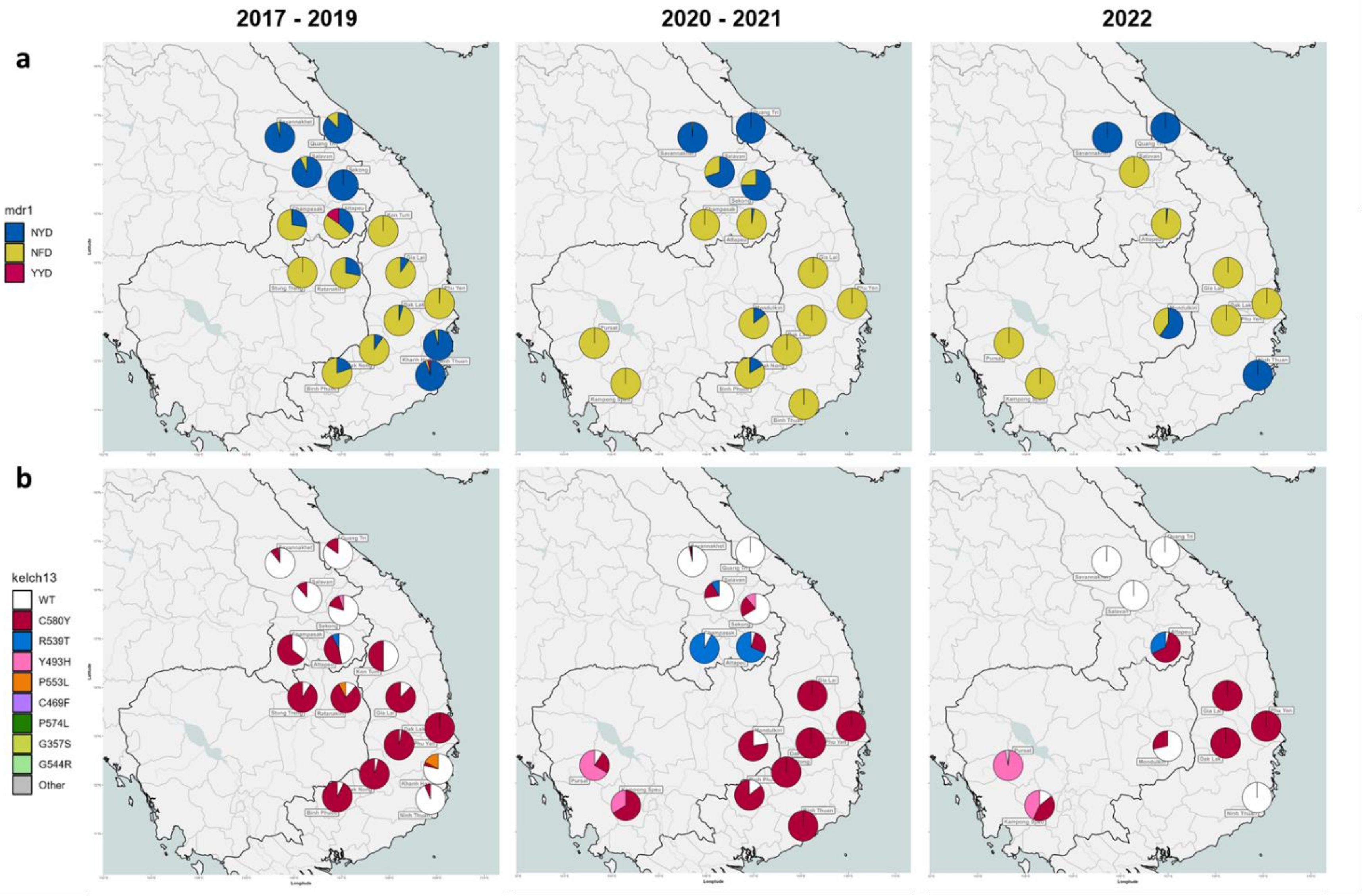
2017 - 2019
2020 - 2021
2022
a
mdr1
NYD
NFD
YYD
b
kelch13
WT
C580Y
R539T
Y493H
P553L
C469F
P574L
G357S
G544R
Other

### Supplementary Methods

**Sample genotyping**

DNA was extracted from DBS samples and processed using the SpotMalaria v2 amplicon sequencing and genotyping platform. Details of the sample processing procedures, and the set of amplicons and variants targetted, are given in the "SPOTmalaria Technical Notes and Methods", available at https://www.malariagen.net/resource/29/, released as supplementary material for Jacob CG *et al.*, 2021.[14] The FASTQ files produced by sequencing were then processed by an informatics pipeline that genotyped the samples and produced the Genetic Report Cards datasets that were used in the analyses. The pipeline is available open-source at https://github.com/genomic-surveillance/AmpRecon with documentation, including a complete workflow. Here, we give an overview of functional blocks that are relevant to the results presented.

**Alignment and Read Counts**. Illumina short read pairs were aligned against reference amplicon sequences, obtained from the 3D7 reference genome V3; reference files are available from https://github.com/genomic-surveillance/AmpReconResources/tree/main/plasmodium/falciparum. Alignments were processed to remove reads that did not meet quality criteria, as detailed in the "SPOTmalaria Technical Notes and Methods", and `bcftools mpileup` (https://samtools.github.io/bcftools/bcftools.html) was used to extract VCF files containing the read depth of each allele at every position of interest.

**Variant Genotyping**. At every position of interest, nucleotide genotypes were established by analyzing allele read depths. If the position was covered by fewer than 10 reads, the genotype was considered "missing" (i.e. undetermined due to insufficient coverage). Otherwise, the sample was deemed as carrying all alleles that were in no less than 5 reads AND in at least 10% of the total reads at that position. If a single nucleotide allele met these criteria, the sample was genotyped as homozygous for that allele; if more than one allele met the criteria, the sample was genotyped as heterozygous. Amino acid alleles were genotyped by concatenating the nucleotide alleles of the adjacent positions in the codon, and then translating the codon to an amino acid. The amino acid genotype was deemed missing if any of the codon's nucleotide genotypes were missing.

**Barcodes**. For each sample, a 101-SNP barcode was called, simply by concatenating the genotypes from each of the positions. Barcode missingness was estimated as the proportion of sites that had a missing genotype; barcode heterozygosity was estimated as the proportion of non-missing positions that carried a heterozygous genotype. Barcode missingness was used as a quality measure, as it increases when low levels of DNA concentration reduce sequencing yield.

***kelch13* genotyping**. Nucleotide positions covering *kelch13* amino acid positions 340-695 were genotyped for each sample, using 6 overlapping amplicons. The genotypes were then scanned to detect changes in the amino acid sequence. If more than 25% of positions was missing, the *kelch13* genotype was deemed "missing". Otherwise, the sample was genotyped as "wild type" ("WT") if all the amino acid positions were called with the same allele as the 3D7 reference. If a single amino acid change caused by a homozygous nucleotide allele was detected, the sample was assigned the relevant mutation. However, if the nucleotide allele that caused the amino acid change was heterozygous, the position was considered heterozygous (i.e. harbouring both mutant and wild type parasites). Similarly, the detection of multiple amino acid changes produced a heterozygous allele.

***plasmepsin 2/3* amplifications**. *Plasmepsin 2/3* (*pm23*) amplifications were determined by combining results from two methods: breakpoint sequence identification[14,12] and qPCR.[15] Copy numbers were not quantified, so the test outcomes were only "wild type" ("WT") if there was a single copy of the gene, or "Amplified" if multiple copies were detected. When both methods produced matching results, or if only one method produced a result, that outcome was assigned as the result. In the presence of discrepancies between the two methods, a "missing" (undetermined) status was assigned.

***mdr1* amplifications**. Amplifications of the *Pf* genes *multidrug resistance 1 transporter* (*mdr1*) was detected by qPCR.[15] Copy numbers were not quantified, so the test outcomes were only "wild type" ("WT") if there was a single copy of the gene, or "Amplified" if multiple copies were detected. A "missing" (undetermined) value was assigned when qPCR could not determine an amplification status.

**Phenotype Predictions**. The amino acid genotypes at specific positions were used to derive predictions of the drug resistance status for a variety of drugs. The prediction rules are detailed in "Phenotype Rules", available at http://ngs.sanger.ac.uk/production/malaria/Resource/29/20200705-GenRe-05-PhenotypeRules-0.39.pdf, released as supplementary material for Jacob CG et al., 2021.[14]

3 World Health Organizaion. Mekong malaria elimination programme: epidemiology summary, volume 12, October-December 2020. (World Health Organisation 2020).

4 World Health Organizaion. Mekong malaria elimination programme: epidemiology summary, volume 20, October–December 2022. (World Health Organisation 2023).

5 World Health Organizaion. Mekong malaria elimination programme: Accelerating malaria elimination in the Greater Mekong, bulletin 10. (2022).

6 National Centre for Parasitology, E. a. M. C. C. Kingdom of Cambodia National Malaria Programme Review 2019. (2019).

7 Srean, C. *et al.* Descriptive assessment of COVID-19 responses and lessons learnt in Cambodia, January 2020 to June 2022. *BMJ Global Health* **8**, e011885 (2023). https://doi.org/10.1136/bmjgh-2023-011885

8 World Health Organization. Lao PDR Coronavirus Disease 2019 (COVID-19) Situation Report #12. (2020).

9 A3M Global Monitoring GmbH.

10 Vietnam Ministry of Health. National Guideline of Malaria Diagnosis and Treatment. (Hanoi, 2020).

11 Cao, V. A. T. *et al.* Exploring the feasibility of introducing triple artemisinin-based combination therapy in the malaria treatment policy in Vietnam. *Malaria Journal* **22**, 326 (2023). https://doi.org/10.1186/s12936-023-04763-4

12 Wikipedia contributors. Vietnamese government response to the COVID-19 pandemic. (2025). <https://en.wikipedia.org/w/index.php?title=Vietnamese_government_response_to_the_COVID-19_pandemic&oldid=1271927022>.

13 Wikipedia. *Vietnamese government response to the COVID-19 pandemic*.

14 Jacob, C. G. *et al.* Genetic surveillance in the Greater Mekong subregion and South Asia to support malaria control and elimination. *Elife* **10** (2021). https://doi.org/10.7554/eLife.62997

15 Ansbro, M. R. *et al.* Development of copy number assays for detection and surveillance of piperaquine resistance associated plasmepsin 2/3 copy number variation in Plasmodium falciparum. *Malaria Journal* **19**, 181 (2020). https://doi.org/10.1186/s12936-020-03249-x
